# Cell-Free Tumor DNA Dominant Clone Allele Frequency (DCAF) Is Associated With Poor Outcomes In Advanced Biliary Cancers Treated With Platinum-Based Chemotherapy

**DOI:** 10.1101/2021.11.01.21265773

**Authors:** Pedro Luiz Serrano Uson Junior, Umair Majeed, Jun Yin, Gehan Botrus, Mohamad Bassam Sonbol, Daniel H. Ahn, Jason S. Starr, Jeremy C Jones, Hani Babiker, Samantha R Inabinett, Natasha Wylie, Ashton WR Boyle, Tanios S. Bekaii-Saab, Gregory J Gores, Rory Smoot, Michael Barrett, Bolni Nagalo, Nathalie Meurice, Natalie Elliott, Joachim Petit, Yumei Zhou, Mansi Arora, Chelsae Dumbauld, Oumar Barro, Alexander Baker, James Bogenberger, Kenneth Buetow, Aaron Mansfield, Kabir Mody, Mitesh J. Borad

**Affiliations:** Division of Hematology & Oncology, Department of Medicine, Mayo Clinic, Scottsdale, AZ, USA; Division of Hematology & Oncology, Department of Medicine, Mayo Clinic, Jacksonville, FL, USA; Division of Gastroenterology and Hepatology, Department of Internal Medicine, Rochester, MN, USA; Division of Medical Oncology, Mayo Clinic, Rochester, MN, USA; Center for Individualized Medicine, Mayo Clinic, Rochester, MN, USA; Arizona State University, Tempe, AZ, USA; Department of Molecular Medicine, Rochester, MN, USA; Mayo Clinic Cancer Center, Phoenix, AZ, USA; Division of Clinical Trials and Biostatistics, Mayo Clinic, Rochester, MN, USA; Hospital Israelita Albert Einstein, São Paulo, Brazil; Department of Pathology, University of Arkansas for Medical Sciences, Little Rock, AR, USA

**Keywords:** Biliary tract cancers, cell-free tumor DNA, liquid biopsy, cholangiocarcinoma

## Abstract

**PURPOSE:** This investigation sough to evaluate the prognostic value of pre-treatment ctDNA in metastatic biliary tract cancers (BTC) treated with platinum based first-line chemotherapy treatment.

**METHODS:** We performed a retrospective analysis of 67 patients who underwent ctDNA testing before platinum-based chemotherapy for first-line treatment for metastatic BTC. For analysis we considered the detected gene with highest variant allele frequency (VAF) as the dominant clone allele frequency (DCAF). Results of ctDNA analysis were correlated with patients’ demographics, progression-free survival (PFS) and overall survival (OS).

**RESULTS:** The median age of patients was 67 years (27-90). 54 (80.6%) of 67 patients evaluated had intrahepatic cholangiocarcinoma; seven had extrahepatic cholangiocarcinoma and six gallbladder cancers. 46 (68.6%) of the patients were treated with cisplatin plus gemcitabine, 16.4% of patients received gemcitabine and other platinum (carboplatin or oxaliplatin) combinations while 15% of patients were treated on a clinical trial with gemcitabine and cisplatin plus additional agents (CX4945, PEGPH20 or nab-paclitaxel). TP53, KRAS, FGFR2, ARID1A, STK11 and IDH1 were the genes with highest frequency as DCAF. Median DCAF was 3% (0-97%). DCAF >3% was associated with worse OS (median OS: 10.8 vs. 18.8 months, p=0.032). Stratifying DCAF in quartiles, DCAF>10% was significantly related to worse PFS (median PFS: 3 months, p=0.014) and worse OS (median OS: 7.0 months, p=0.001). Each 1% increase in ctDNA was associated with a hazard ratio of 13.1 in OS when adjusting for subtypes, metastatic sites, size of largest tumor, age, sex, and CA19-9.

**CONCLUSION:** DCAF at diagnosis of advanced BTC can stratify patients who have worse outcomes when treated with upfront platinum-based chemotherapy. Each increase in %ctDNA decrease survival probabilities.

## INTRODUCTION

Biliary tract cancers (BTCs) include intrahepatic cholangiocarcinoma (IHC), gallbladder cancer (GBC) and extrahepatic cholangiocarcinoma (EHC) and ampulla of Vater cancers (AVC) [1]. BTC represents 3% of gastrointestinal malignancies with 11,980 cases expected to be diagnosed in 2021 [2, 3]. As BTC usually present at an advanced stage only 20% of these tumors are considered resectable [4]. In patients with unresectable disease the 5-year overall survival is about 4% [5].

The survival gain with first-line chemotherapy regimens in BTC is modest since most patients develop progressive disease with a median overall survival (OS) of less than a year [6]. This has generated interest in using next-generation tumor genomic profiling (NGS) and liquid tumor biopsy on peripheral blood to look for targetable genetic alterations [7, 8].

Circulating tumor DNA (ctDNA) has been shown to carry tumor-specific genetic or epigenetic alterations including point mutations, copy number variations, chromosomal rearrangements, and DNA methylation. This ctDNA is released into the circulation after tumor cells undergo apoptosis or necrosis [9]. Evaluation of ctDNA can identify patient specific tumoral genetic alterations while allowing for serial monitoring of tumor genomes in a non-invasive and accurate manner [8]. Therapeutically relevant alterations were seen in ctDNA in 55% of biliary tract cancer patients [8]. Due to these findings the strategy is being used in the setting of advanced disease for treatment selection [10]. Furthermore, it has also been used as an early marker of response to treatment and to track mechanisms of acquired resistance [11].

In colon and breast cancer ctDNA has been used to predict response to treatment and prognosis in the adjuvant and neoadjuvant setting respectively [12, 13]. One marker of interest is variant allele frequency (VAF), which is the number of mutant molecules over total number of wild-type molecules at a specific location on the genome. Pairawan et.al showed that VAF is a surrogate marker of tumor burden and maximum VAF (VAFmax) correlated negatively with prognosis and survival in metastatic cancer [14].

In this study we hypothesized that the dominant clone allele frequency (DCAF) on ctDNA in biliary tract cancer would be associated with overall survival (OS) and progression free survival (PFS) and can serve as a surrogate of disease volume and severity. In addition, we looked at relationship of DCAF to treatment response with first-line platinum-based chemotherapy and clinical demographics.

## MATERIALS AND METHODS

### Patients

From July 2016 through June 2020, 67 patients with advanced BTC underwent ctDNA testing at diagnosis using an available assay [Guardant Health, Inc. (Redwood City, CA)]. All the patients received care at Mayo Clinic Cancer Center in Arizona and Florida. The analysis from this cohort was reviewed and approved by the Mayo Clinic institutional review board. Clinical and demographic information of all patients are included in **Table 1**.

**Table 1:** Demographic characteristics.

|  | Overall (N=67) |
| --- | --- |
| **Age at diagnosis, years** |  |
| Median (range) | 67 (27, 90) |
| **Sex** |  |
| Male | 25 (37.4%) |
| Female | 42 (62.6%) |
| **Diagnosis** |  |
| Extrahepatic cholangiocarcinoma | 7 (10.4%) |
| Gallbladder carcinoma | 6 (9.0%) |
| Intrahepatic cholangiocarcinoma | 54 (80.6%) |
| **Grouping by genomic alterations** |  |
| FGFR2 | 9 (13.4%) |
| ERBB2 | 6 (9.0%) |
| IDH1 | 11 (16.4%) |
| KRAS | 9 (13.4%) |
| Not detectable | 1 (1.5%) |
| Other (TP53, CDKN2A, BRAF, STK11, APC, RET, ARID1A, EGFR, ATM) | 31 (46.3%) |
| **Treatment** |  |
| Gemcitabine + Cisplatin | 46 (68.6%) |
| Gemcitabine + Cisplatin + CX-4945 | 6 (8.95%) |
| Gemcitabine + Oxaliplatin | 6 (8.95%) |
| Gemcitabine + Carboplatin | 5 (7.5%) |
| Gemcitabine + Cisplatin + Nab-paclitaxel | 3 (4.5%) |
| Gemcitabine + Cisplatin + PEGPH20 | 1 (1.5%) |
| **Size of largest lesion (cm)** |  |
| Median (range) | 6 (2-19) |
| **Metastatic sites** |  |
| Liver | 24 (35.8%) |
| Extrahepatic only (ex. lung, bones, peritoneum, lymph nodes) | 4 (6%) |
| Liver + extrahepatic | 39 (58.2%) |
| **Pre-Treatment CA19-9 Level (U/mL)** |  |
| Median (range) | 103 (1-103198) |
| **Pre-Treatment Maximum Mutant Allele Frequency (%)** |  |
| Mean CI | 10 (6, 14) |
| Median (range) | 3 (0 – 94) |
| Q1, Q3 | 1, 10 |
| **Tumor response (partial response + complete response)** |  |
| No | 34 (57.6%) |
| Yes | 25 (42.4%) |
| **Disease Control (partial response + complete response + stable disease)** |  |
| No | 21 (35.6%) |
| Yes | 38 (64.4%) |
| **Progression-free survival events** |  |
| No | 31 (46.3%) |
| Yes | 36 (53.7%) |
| **Overall survival events** |  |
| No | 28 (41.8%) |
| Yes | 39 (58.2%) |

### Comprehensive genomic testing in plasma

Circulating tumor DNA was extracted from whole blood. ctDNA fragments, both leukocyte- and tumor-derived, were simultaneously sequenced. The VAF was calculated as the proportion of ctDNA harboring the variant in a background of wild type ctDNA. Analytical information, bioinformatics analysis and Guardant360 database has been previously described [15, 16].

### Outcomes

Assessments regarding response to therapy (complete response [CR], partial response [PR], stable disease [SD], disease progression [PD]) were retrospectively collected by review of patient’s charts. Positive response to therapy was considered PR and CR by Response Evaluation Criteria in Solid Tumors (RECIST). Disease control rate was determined based on CR, PR, and SD. Progression-free survival (PFS) was determined by the time during treatment with chemotherapy and after without disease progression. Overall survival (OS) was determined by the time of diagnosis of advanced disease till death or last day of follow up for patients on treatment and alive.

### Statistical analysis

We summarized categorical data as frequency counts and percentages, and continuous measures as means, standard deviations, medians, and ranges. Categorical variables were compared using the chi-square test or Fisher’s exact test. Continuous variables were compared using the one-way ANOVA test or Kruskal–Wallis test. Multivariate logistic regressions were performed to assess the association of ctDNA with response rate and disease control rate with adjustment for disease subtype, age, sex, CA19-9, lesion size, and metastatic site. The distributions of time-to-event outcomes were estimated using the Kaplan-Meier methods and compared between low versus high ctDNA dichotomized by the median dominant clone allele frequency (i.e., low < 3% versus high >= 3% ctDNA) using the log-rank test. Hazard ratios (HRs) and 95% confidence intervals (CIs) were estimated using a multivariate Cox model adjusting for disease subtype, age, sex, CA19-9, lesion size, and metastatic site. Sensitivity analysis were conducted to explore either 3-quantiles (<= 33% percentile, 34-66% percentile, > 66% percentile) or quartiles as the cutoffs in dominant clone allele frequency.

## RESULTS

### Patient demographics

A total of 67 patients were included in the analysis. 80.6% (54) had intrahepatic cholangiocarcinoma, 10.4% (7) patients had extrahepatic cholangiocarcinoma and 9% (6) had gallbladder cancer. All patients included had ctDNA collected before the first-line chemotherapy regimen for advanced disease. Median age of all patients was 67 y/o (27-90) and the majority were female (62.6%). All patients were treated with platinum-based chemotherapy regimens as first-line treatment. Most patients (68.6%) were treated with cisplatin plus gemcitabine, eleven (16,4%) patients received gemcitabine plus other platinum (carboplatin or oxaliplatin) combinations while ten (15%) patients were treated on a clinical trial with gemcitabine and cisplatin plus additional agents (CX4945, PEGPH20 or nab-paclitaxel). The median size of largest lesion was 6 cm (2-19cm) and more than half (58.2%) had multiple metastatic sites including liver and extrahepatic sites. Lungs, bones, lymph nodes and peritoneum were the sites with most extrahepatic metastasis identified. Other clinical information can be found on **Table 1**.

Several potential targetable genes were detected with ctDNA including FGFR2, HER2, IDH, MET, EGFR, BRAF and KRAS. A higher prevalence of TP53 were observed among the three subtypes. Homologous recombinant repair genes were identified in IHC and EHC including ATM and BRCA2. Prevalence of all genomic alterations accordingly to primary tumor can be found on **Figure 1**.

**Figure 1:**
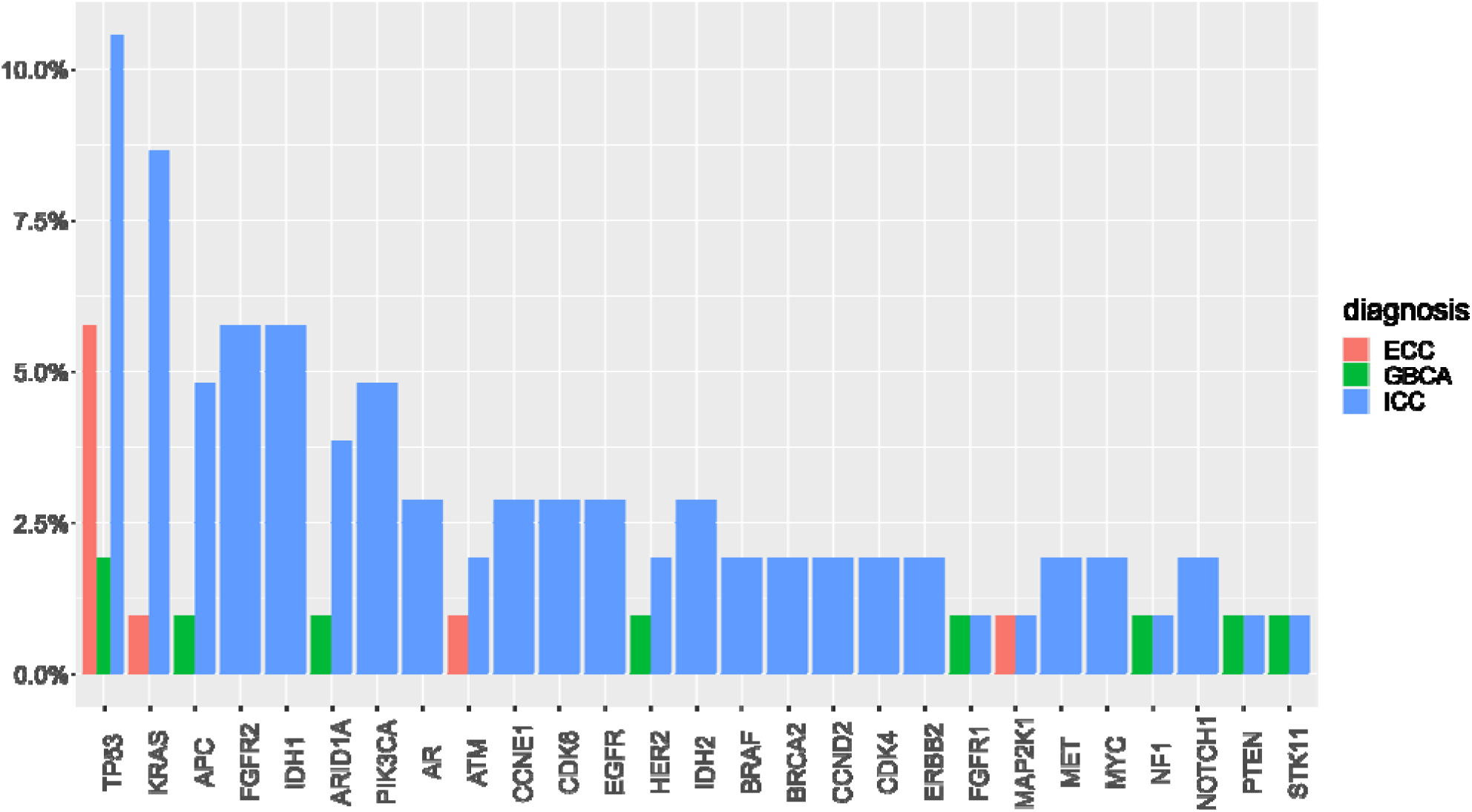
Prevalence of genomic alterations corresponding to primary tumor. ECC: extra hepatic cholangio carcinoma, GBC A: gallbladder carcinoma, ICC: intrahepatic cholangiocarcinoma.

TP53, KRAS, FGFR2, ARID1A, STK11 and IDH1 were the genes with highest variant allele frequency as dominant clone (**Figure 2**). Most ERBB2 (HER-2) genomic alterations detected were amplifications, identified in 4 patients. Other genes with detected amplifications included KRAS, EGFR, BRAF, MET, CCNE1, CCND1, CCND2, MYC, FGFR1, FGFR2, CDK4, CDK6, PIK3CA, AR. For analysis we considered the detected genomic alteration with the highest variant allele frequency (VAF) as the dominant clone allele frequency (DCAF).

**Figure 2:**
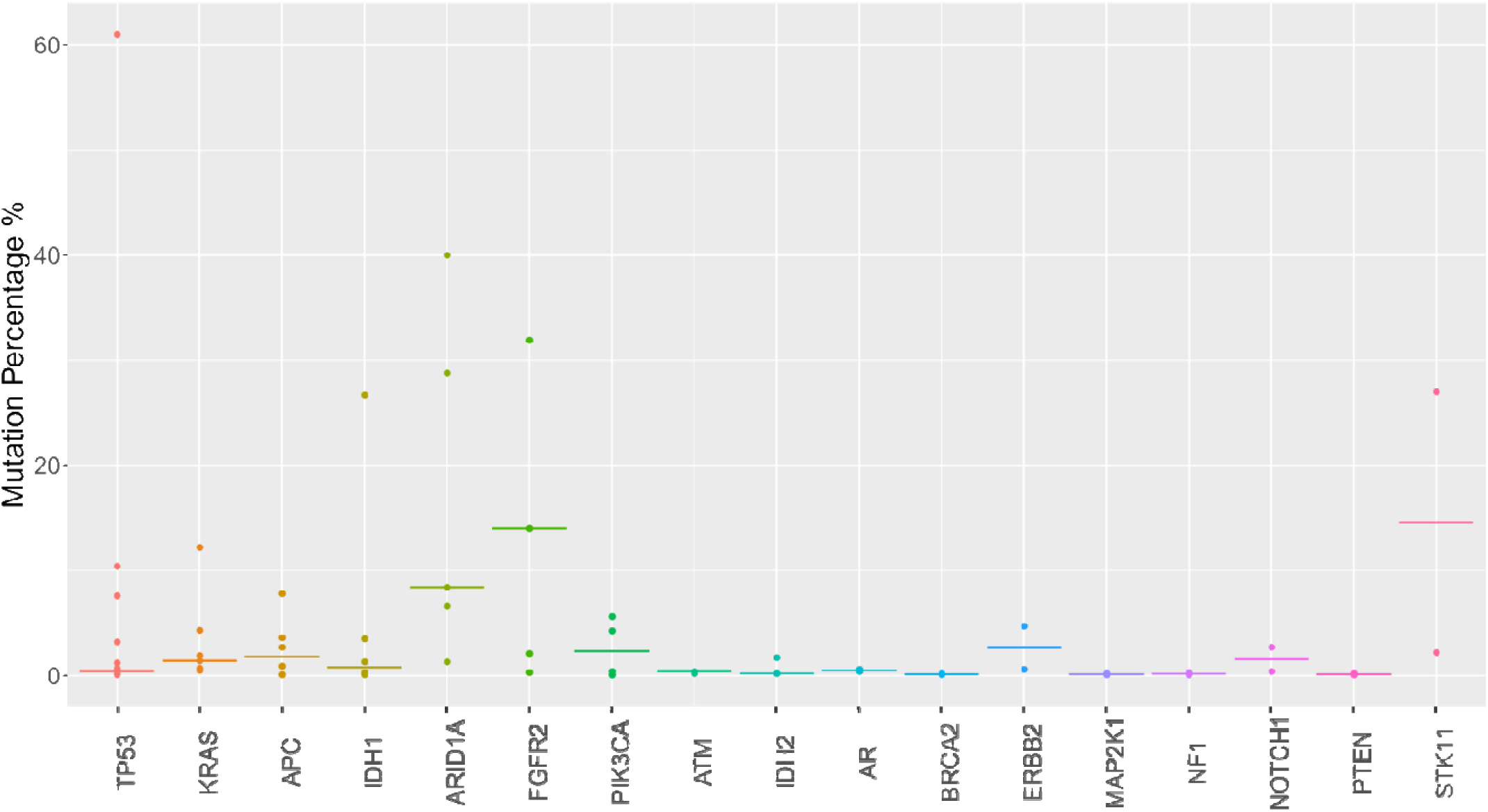
Variant allele frequency of detected genes.

### Dominant clone allele frequency and prognostic factors

One patient with no tumor genomic alteration detected was excluded from this analysis. The median dominant clone allele frequency (DCAF) was 3% (0-97%). DCAF >3% was associated with inferior PFS (median PFS: 4.7 vs. 7.7 months, p=0.087. **Supplementary figure 1**) and significantly worse OS (median OS: 10.8 vs. 18.8 months, p=0.032. **Figure 3**).

**Figure 3:**
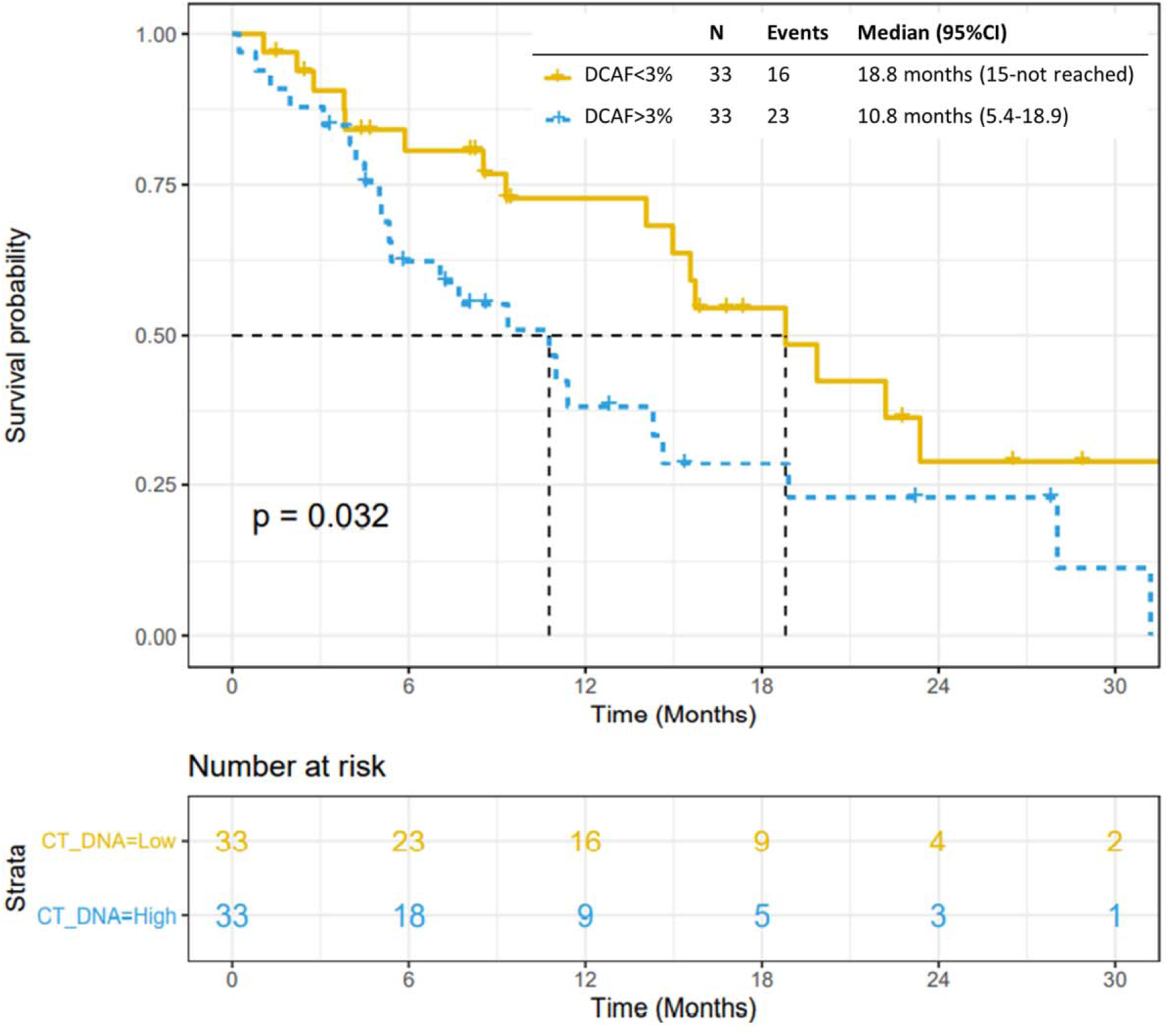
Overall survival Kaplan-Meier curve by DCAF>3%.

We further analyzed DCAF using either 3-quantiles or quartiles as the cutoffs. DCAF divided by 3 quantiles (Q1: ctDNA ≤1%, Q2: ctDNA 1-7%, Q3: ctDNA ≥7%) was significantly associated with PFS (p=0.022) (**Supplementary figure 2)** but not OS (p=0.065) (**Supplementary figure 3**). DCAF divided by quartiles (Q1: ctDNA ≤0.6%, ctDNA Q2: 0.6-3%, ctDNA Q3: 3-10%, ctDNA Q4: ≥10%) was significantly associated with PFS (p=0.014) (**Figure 4**) and OS (p=0.001) (**Figure 5**).

**Figure 4:**
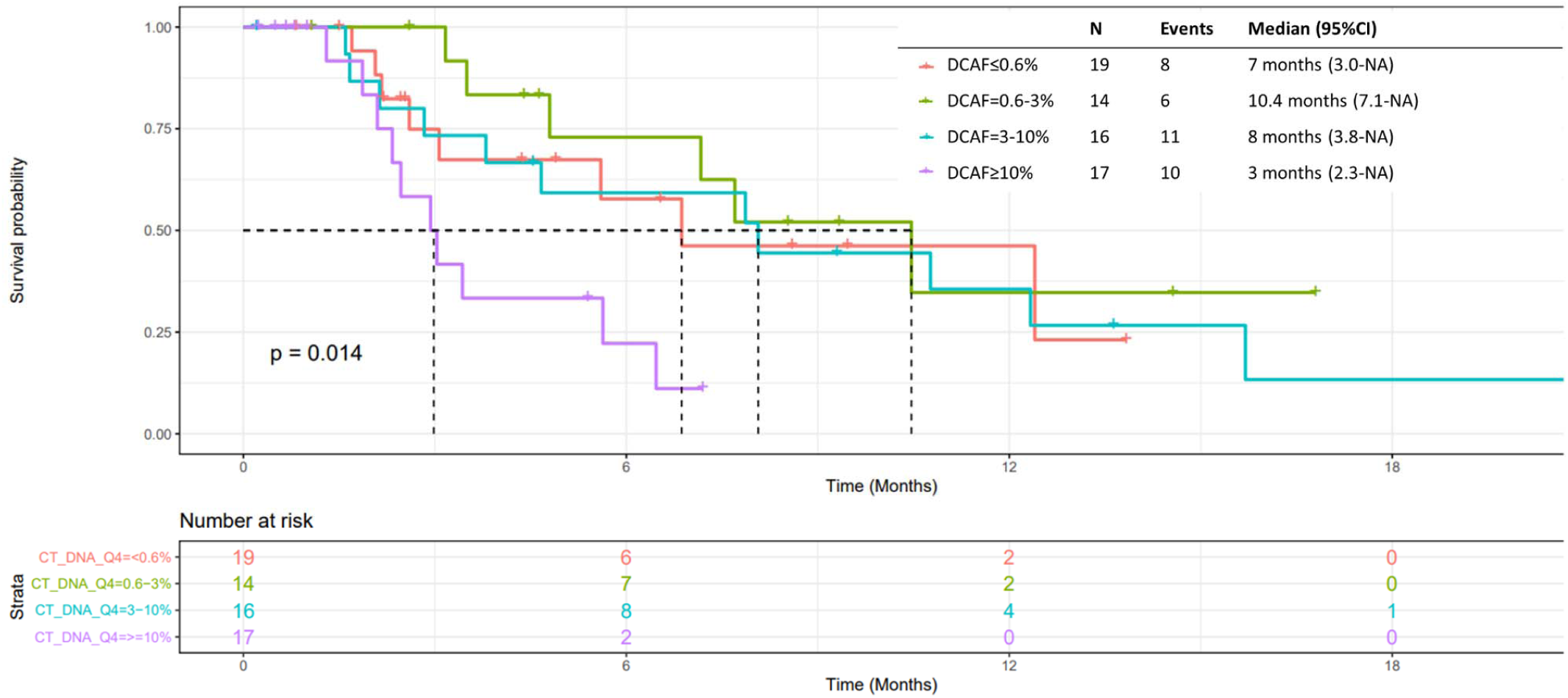
Progression-free survival Kaplan-Meier curve by ctDNA.

**Figure 5:**
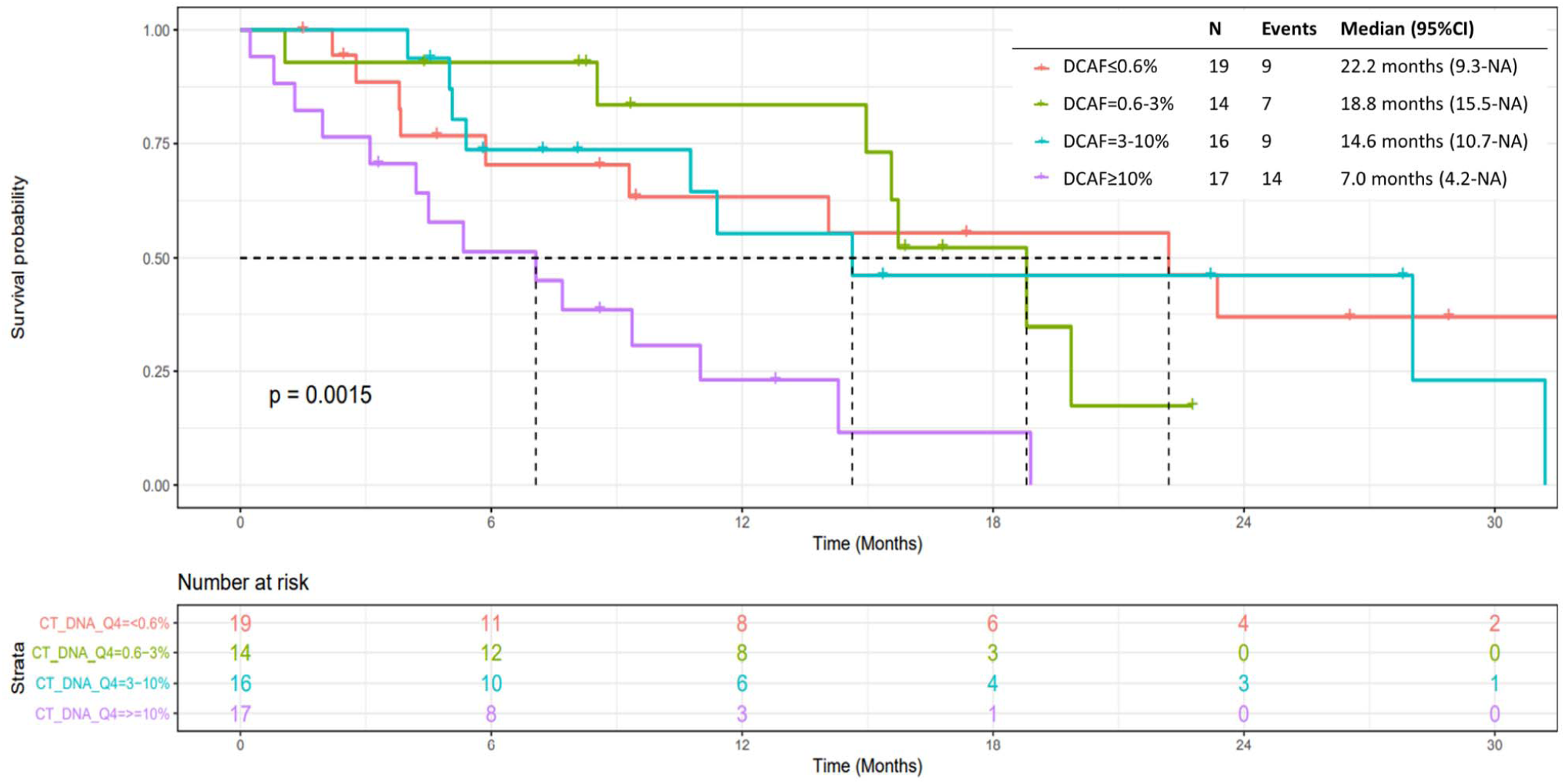
Overall survival Kaplan-Meier curve by groups.

Each 1% increase in ctDNA is associated with a hazard ratio of 13.1 in OS when adjusting for primary tumor, size of the largest lesion, metastatic sites, sex, age, and CA19-9 (**Table 2**). No significant differences in response or disease control rate to chemotherapy was observed in patients with low or high ctDNA (**Supplementary figure 4 and 5**). No statistical significance was found between DCAF and the presence of potential actionable targets including FGFR2, IDH1/2, ERBB2 and KRAS (**Supplementary figure 6)**.

**Table 2:** Multivariate analysis for overall survival.

|  | Hazard Ratio | 95%CI | P value |
| --- | --- | --- | --- |
| Location of primary: non-IHC vs. IHC | 1.54 | [ 0.46 , 5.21 ] | 0.484825 |
| Age at diagnosis, 10yrs | 1.18 | [ 0.83 , 1.67 ] | 0.359703 |
| Pre-Treatment CA19-9 Level ($\geq 100$ U/mL) | 2.08 | [ 0.96 , 4.53 ] | 0.063824 |
| ctDNA (%) | 13.07 | [ 1.2 , 142.32 ] | 0.034866 |
| Sex | 1.24 | [ 0.57 , 2.72 ] | 0.588142 |
| Lesion Size | 1.02 | [ 0.92 , 1.13 ] | 0.762273 |
| Met Site: Extrahepatic (Ref: Liver) | 0.66 | [ 0.08 , 5.47 ] | 0.700660 |
| Met Site: Liver + Extrahepatic (Ref: Liver) | 1.09 | [ 0.49 , 2.43 ] | 0.831125 |

Legend: IHC: intrahepatic cholangiocarcinoma

The interaction between CA19-9 and DCAF were not statistically significant (OS: P_interaction_ = 0.12; PFS: P_interaction_ = 0.06). Although CCA patients with high DCAF and high CA19-9 (DFCA >= 3%, CA19-9 >= 100) had worst OS, no statistical significance was found for PFS (p=0.19) or OS (p=0.13) (**Supplementary figures 7 and 8**).

## DISCUSSION

In this study, we assessed whether the highest variant allele frequency detected by ctDNA, namely DCAF, could be a prognostic factor in patients with advanced biliary tract cancer at diagnosis. Based on the findings, patients with DCAF>3% at diagnosis had worse overall survival when treated with upfront platinum-based chemotherapy. Furthermore, DCAF>10% was significantly related to worse PFS and OS. However, no differences in response rate were observed among patients with DCAF high or low. Moreover, ctDNA proved to be an independent factor related to overall survival in multivariate analysis. Collectively, these data suggest a prognostic and not predictive role for DCAF in patients with advanced biliary tract cancer undergoing platinum-based therapy.

The landscape of ctDNA genomic alterations of biliary tract cancers has already been previously described [8, 17]. Similarly, to our findings, these studies included more patients with IHC, and the genes with the highest detection with ctDNA included principally KRAS, TP53, FGFR2, IDH1 and ARID1A [8, 17]. In our cohort, we observed different patterns of prevalence, with ATM and MAP2K1 detected in extrahepatic cholangiocarcinoma and ERBB2, NF1 and PTEN in gallbladder cancer.

Variant allele frequency is related to outcomes, it is more prominent in metastatic disease and is associated with tumor volume [14, 16, 18]. In metastatic pancreatic cancer, detectable ctDNA and high VAF was associated with worse overall survival [18, 19, 20]. Prognostic significance was observed in other solid tumors including colorectal cancer, breast cancer and prostate cancer [21-23]. Little is known about VAF and prognosis in biliary tract cancers. Lower values of VAF were associated with prolonged progression-free survival in a cohort of 24 patients with cholangiocarcinoma [18]. Considering the highest VAF value, we showed that the DCAF>3% is related to numerically inferior PFS (but not statistically significant) and worse OS in patients with biliary tract cancers treated with standard upfront platinum-based chemotherapy for advanced disease. Interestingly, the DCAF was determined by multiple different genes among the cases, including TP53, KRAS, FGFR2, ARID1A, STK11 and IDH1, suggesting as previously stated by other colleagues that the highest VAF would be a surrogate of disease burden not related specifically to the gene detected. In agreement with this, evaluating the presence of specific genes of interest in the ctDNA overall analysis we did not found any association with DCAF and possible targetable genes including FGFR2, ERBB2, IDH1 and KRAS.

Prognostic factors related to PFS and OS in advanced biliary tract cancers were evaluated from the ABC-02 trial and an international dataset [24]. In this analysis, the authors evaluated prognostic factors in a combined sample size of more than one thousand patients [24]. Although the results suggest multiples factors in multivariate analysis including hemoglobin, gender and neutrophils, receiver operator curve (ROC) analysis suggested that the model generated had a limited prognostic value [24]. Even the primary tumor site was not significant, in contrast to the findings of other groups [25]. After multiple efforts evaluating scores and factors to prognostication of advanced biliary tract cancers [26, 27, 28], the ability to predict prognosis need improvement. In our analysis, the overall survival impact of ctDNA was observed after stratifying with other possible prognostic factors including size of largest lesion, locally advanced/metastatic designation, primary tumor location, metastatic sites, gender, CA 19-9 and age. Evaluating ctDNA as a continuous variable, higher values are related to inferior survival probabilities. Based on the findings, ctDNA and DCAF could be a reliable easy to collect prognostic instrument in prospective trials.

Considering the investigative field of ctDNA in biliary tract cancers, larger, multi-centered prospective studies would be necessary to address the application of ctDNA in the multiple disease assessment junctures, considering early diagnosis, minimal residual disease assessment, monitoring in advanced stages during systemic treatment and assessment of mutations that associate with resistance during treatment with targeted therapies. Evaluation of ctDNA and DCAF in metastatic disease would be a tool for genomic profiling in prospective trials, can be a surrogate of disease volume and will assist in an adequate stratification of patients with advanced biliary tract cancer in randomized studies.

Some limitations of this study include the number of patients, limited institution aspect, inherent limitations associated with a targeted gene panel and the retrospective nature of data collection. Furthermore, most of the patients included had tumor arising from the intrahepatic duct. This limitation is shared with studies in biliary tract cancers and efforts should be made to include patients with extrahepatic and gallbladder carcinomas in initiatives of genomic profiling and ctDNA. Even with the limited number of patients, strong association of ctDNA and OS were observed. This study only evaluated patients treated with upfront chemotherapy. Although this is the standard of care to date, multiple trials are evaluating targeted treatments including FGFR2 inhibitors in the first-line therapy for advanced disease and it would be unclear if upfront tyrosine kinase inhibitors would alter the results. On the other hand, as stated before, the presence of targetable genes had no association with DCAF results and impact of ctDNA on overall survival.

## CONCLUSION

ctDNA is a powerful prognostic tool in advanced biliary tract cancers. DCAF at diagnosis of advanced disease who would receive platinum based systemic therapy identifies patients with worse prognosis. ctDNA should be evaluated in prospective trials as a stratification factor for advanced disease.

## Data Availability

All data produced in the present study are available upon reasonable request to the authors

## ACKNOWLEDGEMENTS

The Mayo Clinic Hepatobiliary SPORE (P50CA 210964) funded the statistical analysis for this project. This work was supported by the National Institute of Health (NIH) through a DP2 Award CA195764 (to MJB); National Cancer Institute (NCI) K12 award CA090628 (to MJB), K01 award CA234324 (to BN), SPORE Project Award 5P50CA210964-03 (to GJG and MJB), SPORE Supplement Award 3P50CA210964-02S1 (to OB), Mayo Clinic Center for Individualized Medicine (CIM) Precision Cancer Therapeutics Program; and Mayo Clinic Cancer Center. The funders had no role in study design, data collection and analysis, decision to publish or preparation of the manuscript. Its contents are solely the responsibility of the authors and do not necessarily represent the official views of the NIH.

## Tables and figures

**Figure 1: Prevalence of genomic alterations accordingly to primary tumor**

**Figure 2: Variant allele frequency of detected genes**

**Figure 3: Overall survival Kaplan-Meier curve by DCAF>3%**

**Figure 4: Progression-free survival Kaplan-Meier curve by ctDNA**

**Figure 5: Overall survival Kaplan-Meier curve by ctDNA**

**Supplementary figure 1: Progression-free survival and DCAF>3%**

**Supplementary figure 2: Progression-free survival and DCAF quartiles**

**Supplementary figure 3: Overall survival and DCAF quartiles**

**Supplementary figure 4: ctDNA Response vs. Non–Response. p = 0**.**2678**

**Supplementary figure 5: ctDNA Disease Control vs. Disease progression. p = 0**.**0843**

**Supplementary figure 6: ctDNA by Group. p = 0**.**1373**

**Supplementary figure 7: CA19-9 and DCAF impact on PFS**

**Supplementary figure 8: CA19-9 and DCAF impact on OS**

