## Supplementary files for "Cell-Free Tumor DNA Dominant Clone Allele Frequency (DCAF) Is Associated With Poor Outcomes In Advanced Biliary Cancers Treated With Platinum-Based Chemotherapy"

### Slide 1
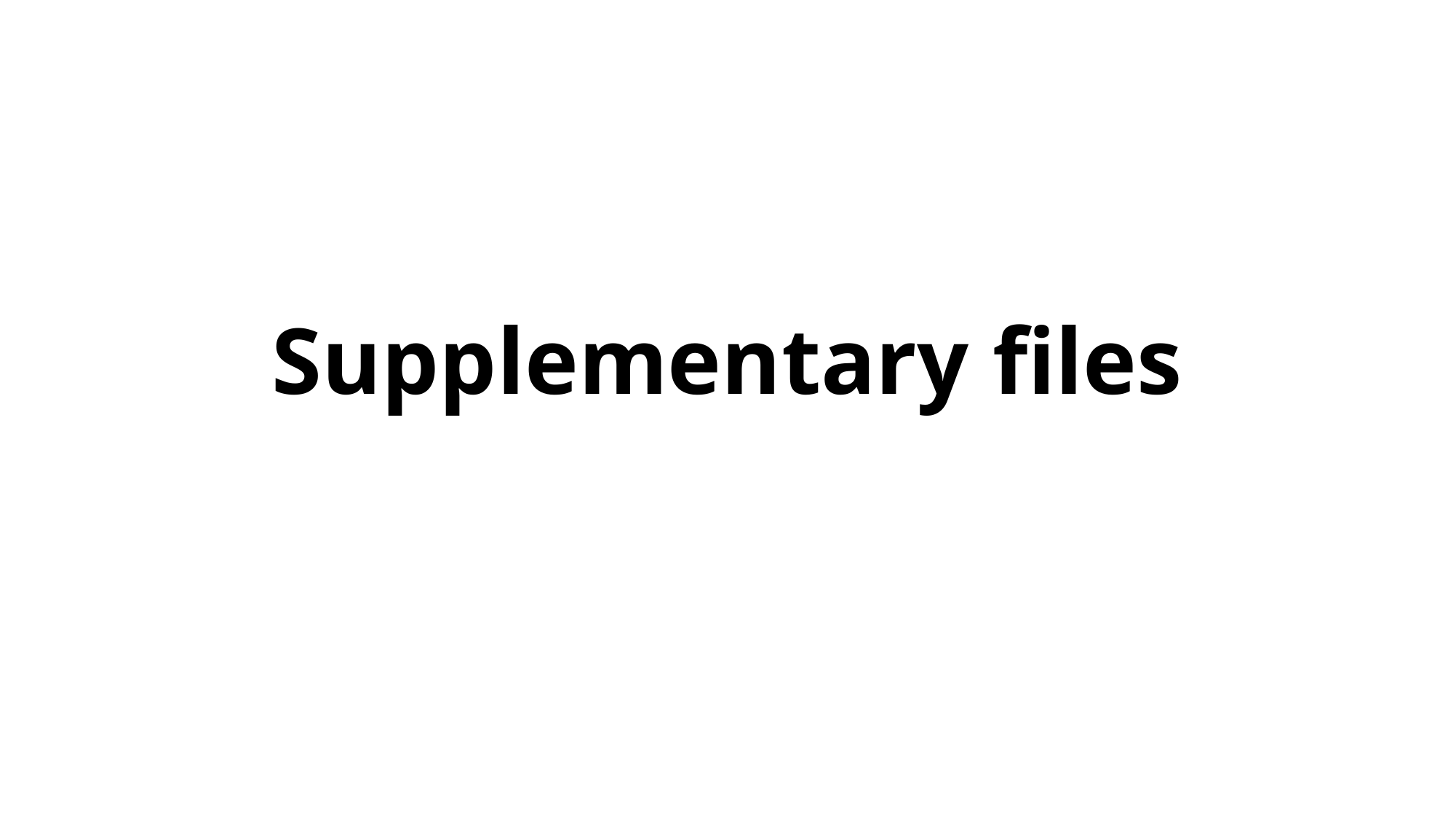

#### Supplementary files

### Slide 2
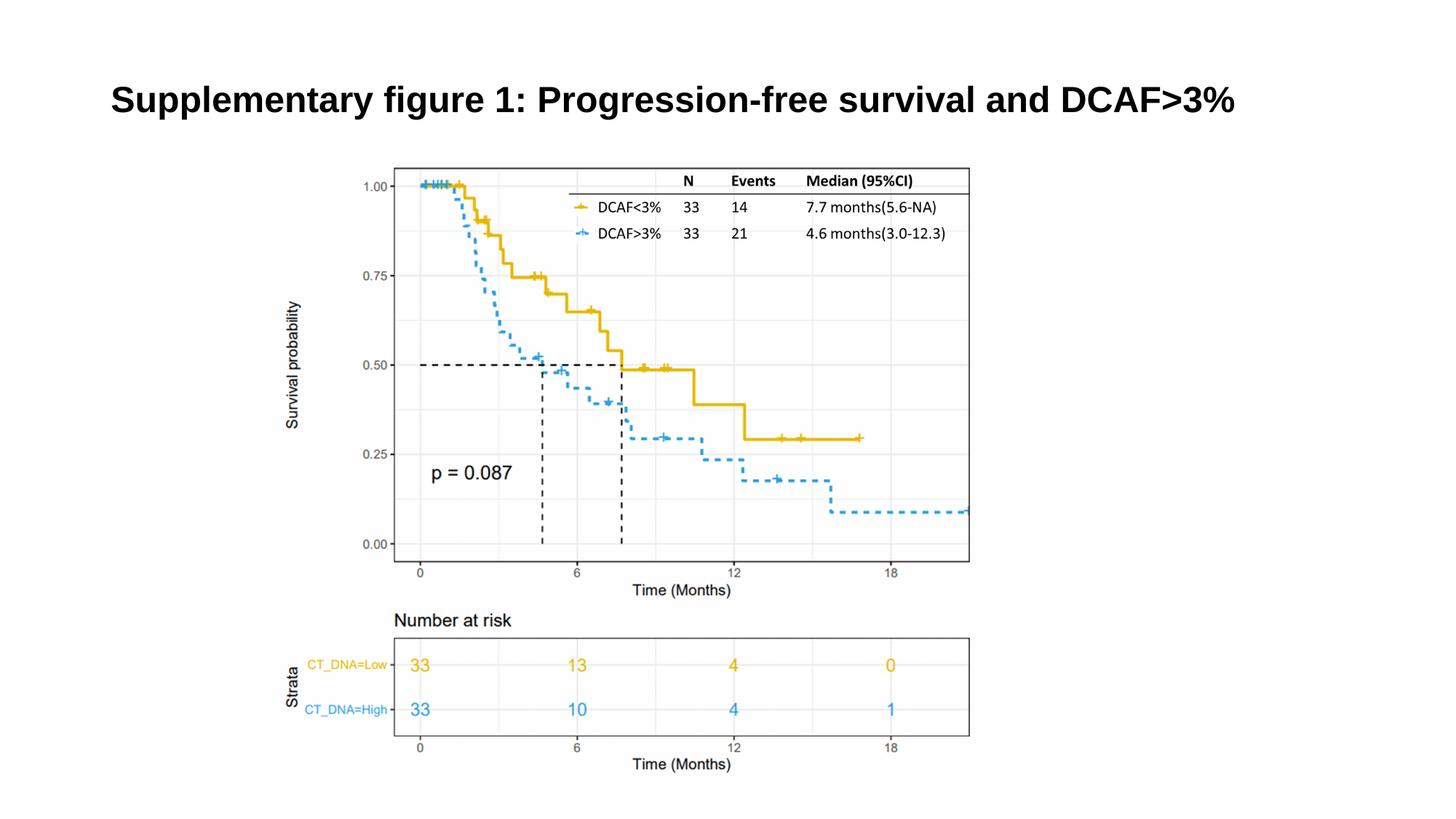

#### Supplementary figure 1: Progression-free survival and DCAF>3%

### Slide 3
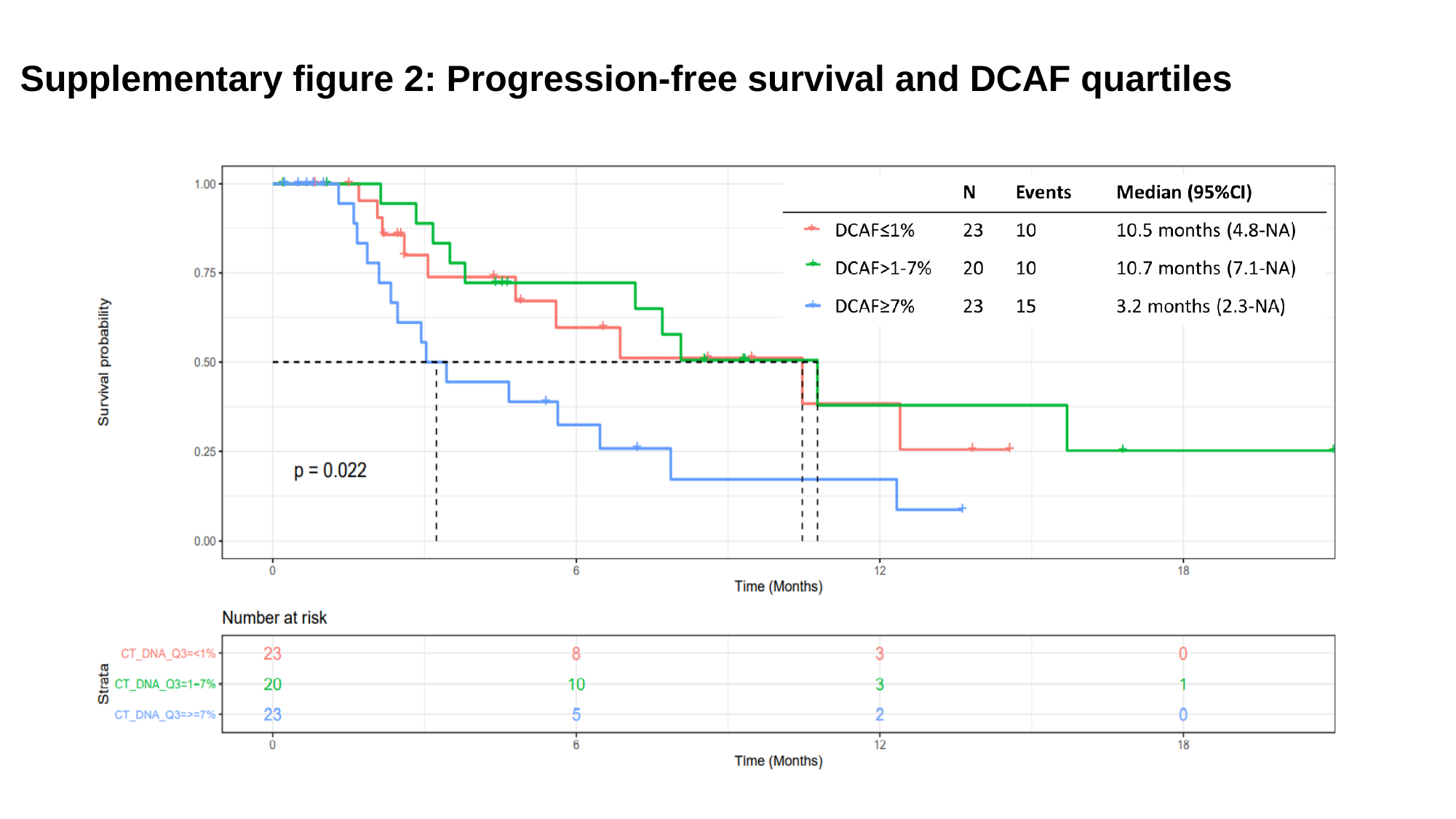

#### Supplementary figure 2: Progression-free survival and DCAF quartiles

### Slide 4
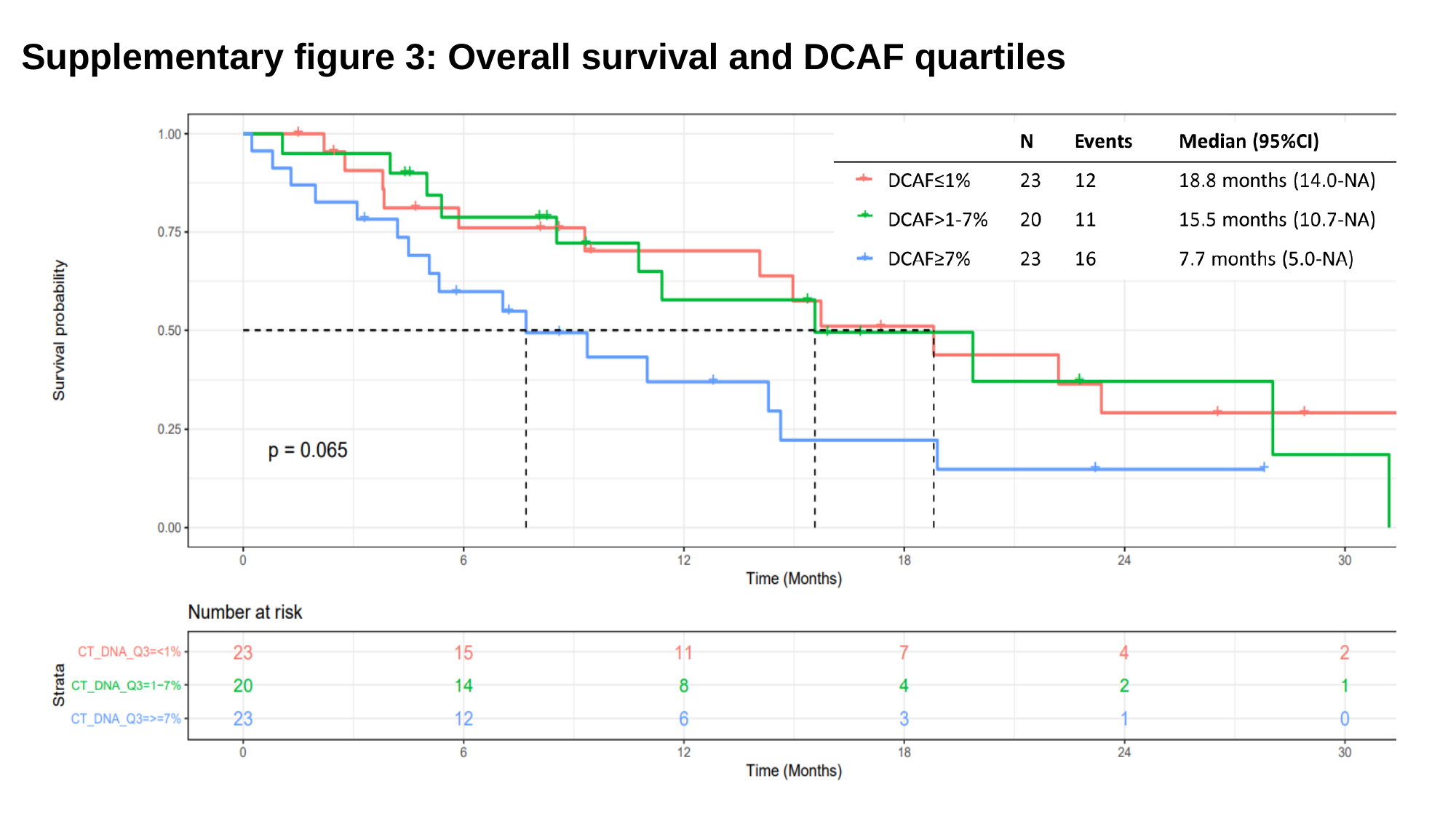

#### Supplementary figure 3: Overall survival and DCAF quartiles

### Slide 5
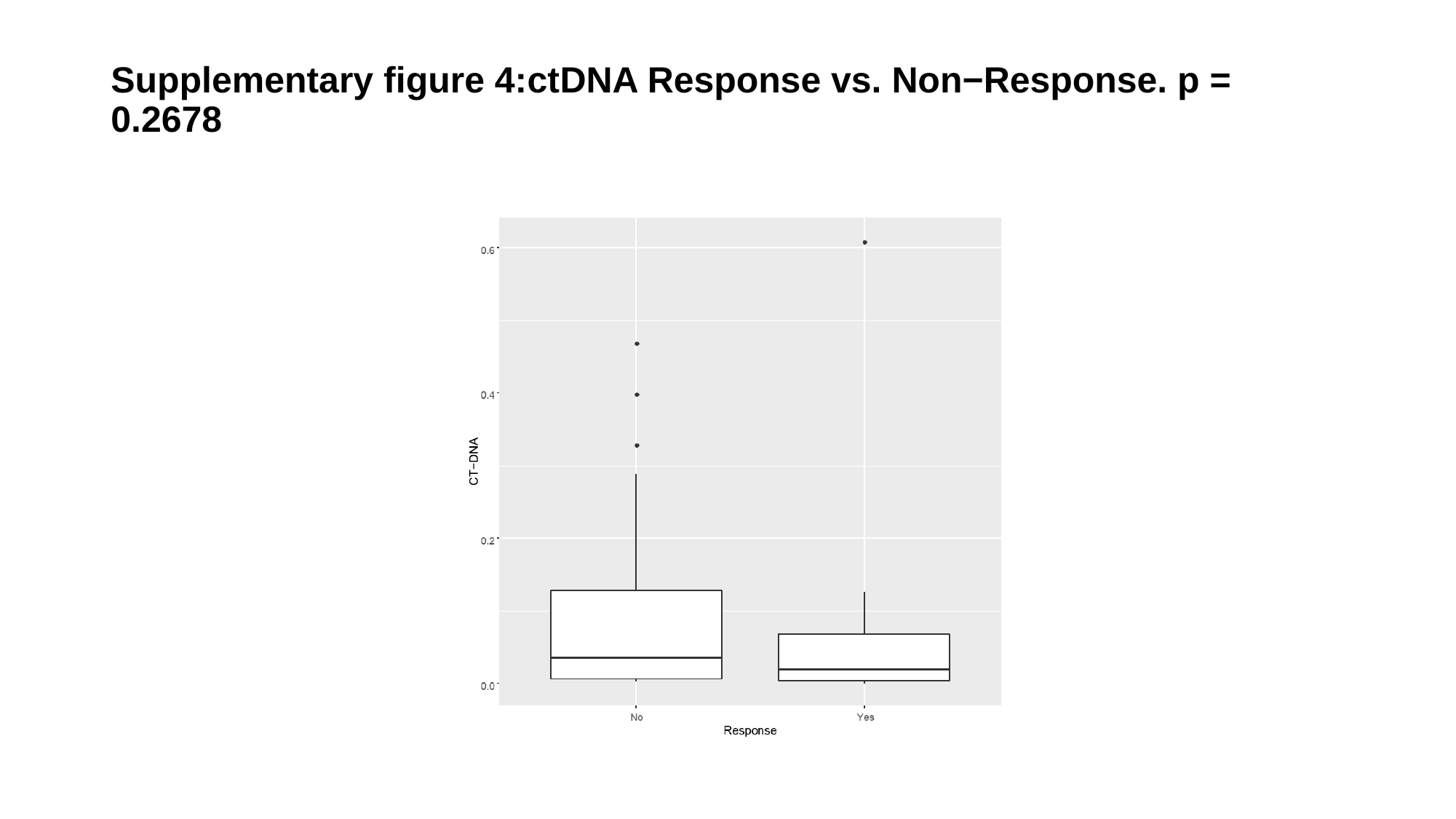

#### Supplementary figure 4:ctDNA Response vs. Non−Response. p = 0.2678

### Slide 6
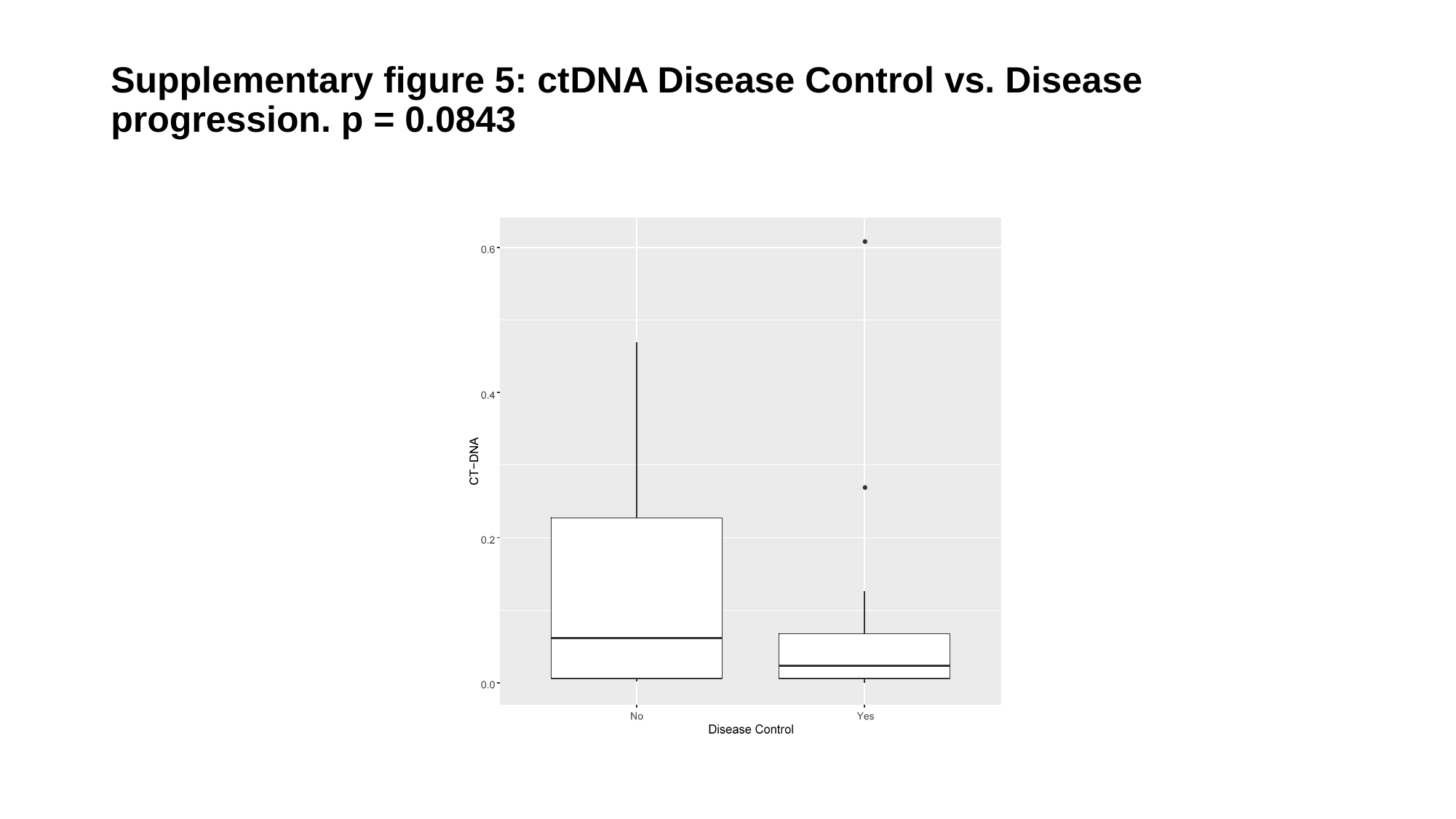

#### Supplementary figure 5: ctDNA Disease Control vs. Disease progression. p = 0.0843

### Slide 7
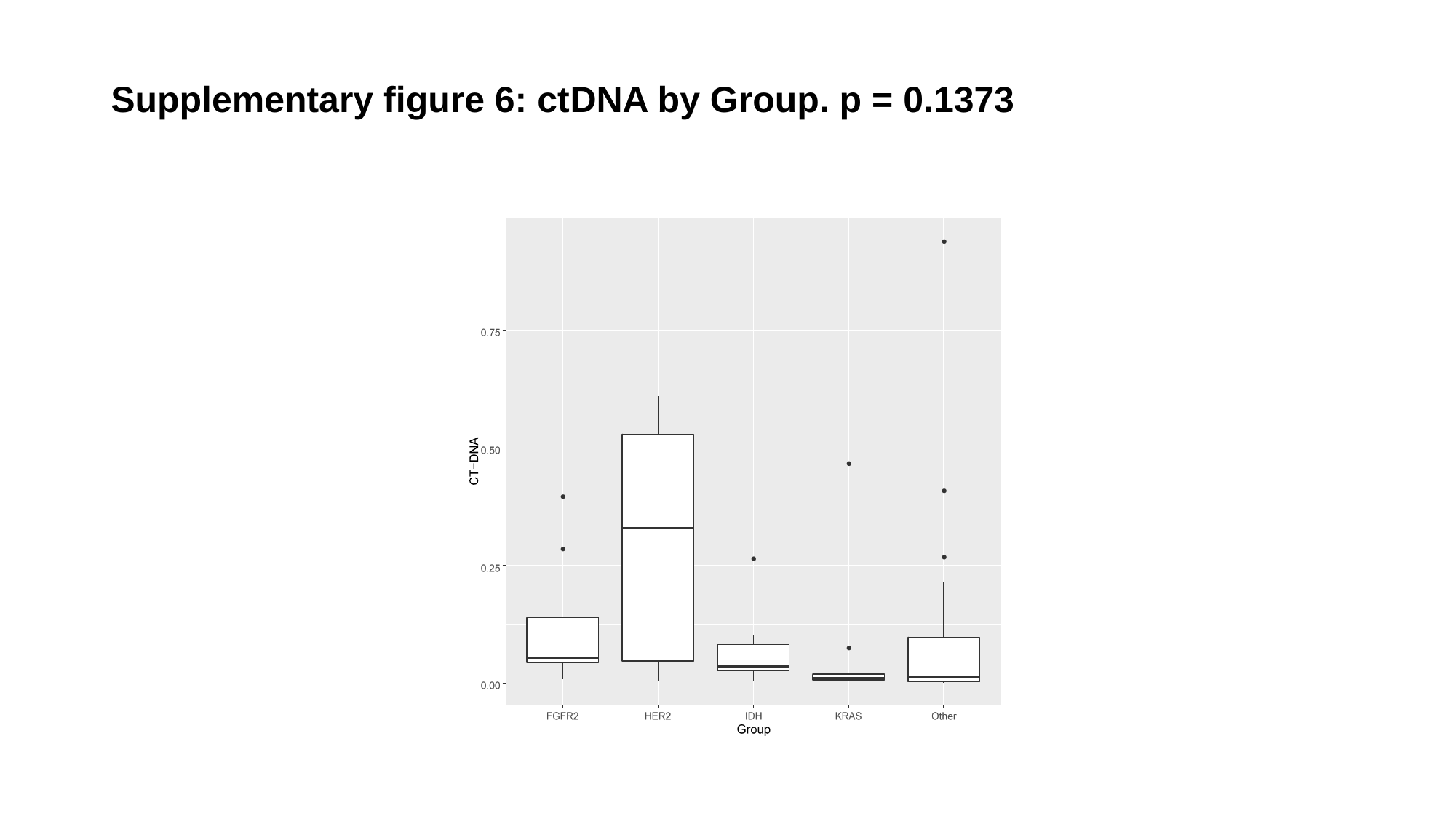

#### Supplementary figure 6: ctDNA by Group. p = 0.1373

### Slide 8
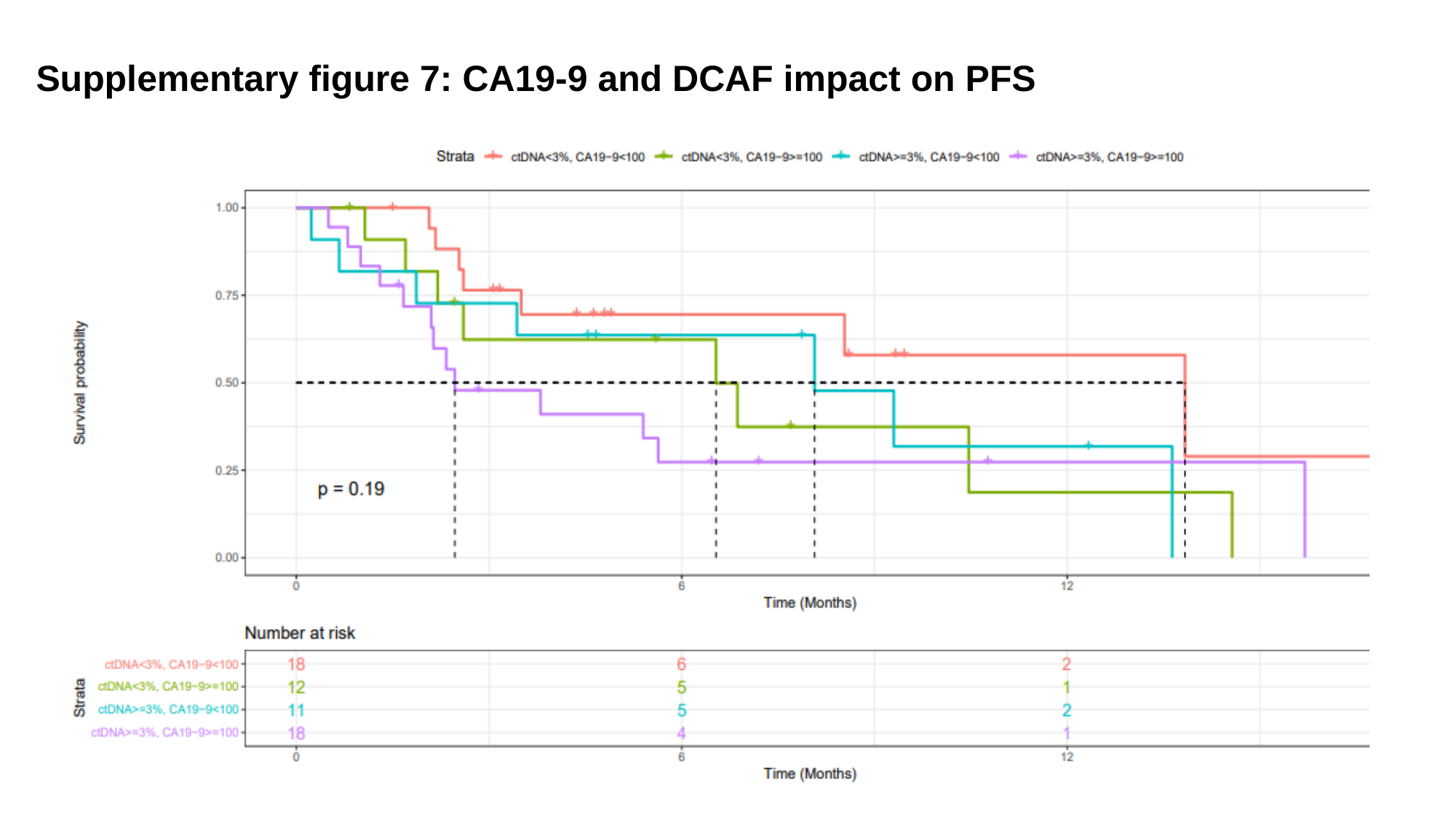

#### Supplementary figure 7: CA19-9 and DCAF impact on PFS

### Slide 9
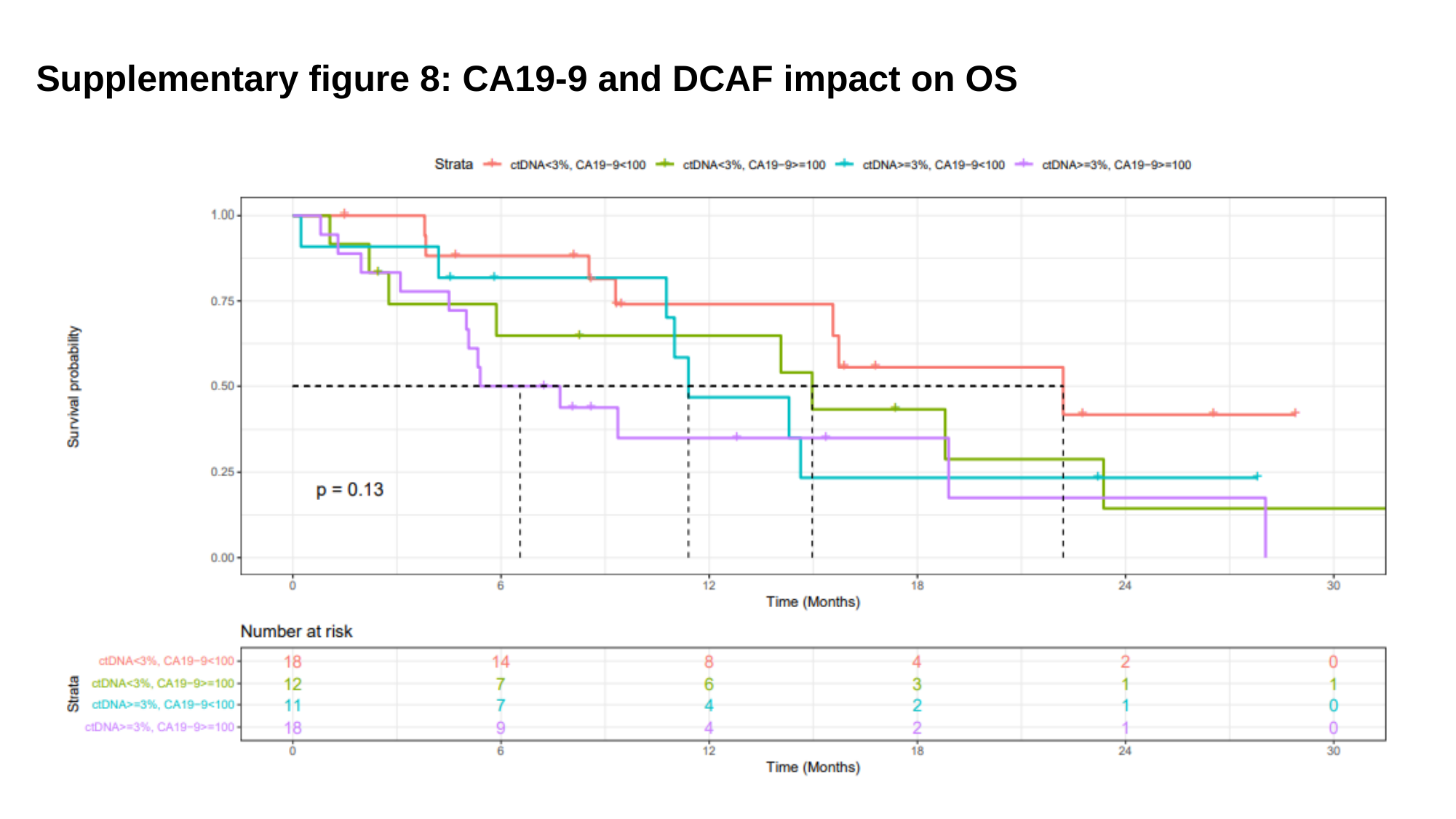

#### Supplementary figure 8: CA19-9 and DCAF impact on OS
